# A nomogram to predict failure rate in immediate implant-based breast reconstruction

**DOI:** 10.1101/2022.06.12.22276288

**Authors:** Meizhen Zhu, Fanrong Zhang, Jiefei Mao, Jun Fang, Daobao Chen

**Affiliations:** Department of Breast Surgery, The Cancer Hospital of the University of Chinese Academy of Sciences (Zhejiang Cancer Hospital), Institute of Basic Medicine and Cancer (IBMC), Chinese Academy of Sciences, Hangzhou, China; Department of Radiation Therapy, The Cancer Hospital of the University of Chinese Academy of Sciences (Zhejiang Cancer Hospital), Institute of Basic Medicine and Cancer (IBMC), Chinese Academy of Sciences, Hangzhou, China

## Abstract

Controversies on risk factors affecting immediate implant-based breast reconstruction (IBBR) still exist. We aimed to evaluate risk factors of reconstruction failure (RF) in immediate IBBR. We retrospectively reviewed 239 patients (241 breasts) who underwent immediate IBBR from 2011 to 2019. A nomogram was established to predict RF rate in stage I operation and logistic regression model was created for stage II operation. 14 (5.81%) and 7 (5.51%) reconstructive breasts experienced RF in stage I and II operation, respectively. Body mass index (BMI) [odds ratio (OR)=1.569, 95% confidence interval (CI): 1.087-2.263, *p*=0.016], implant/tissue expander (TE) volume>86.30% (OR=5.711, 95% CI: 1.067-30.583, *p*=0.042), adjuvant chemotherapy (ACT) (OR=30.094, 95% CI: 2.030-446.084, *p*=0.013) were independent risk factors for RF in stage I operation. The nomogram to predict RF rate in stage I operation revealed area under the curve (AUC) was 0.920 (95% CI: 0.844-0.995). Logistic regression demonstrated that mesh use was the independent predictor of RF in stage II operation (OR=47.326, 95% CI: 3.11-720.081, *p*=0.005). The nomogram exhibited satisfied predictive ability. Present findings enhanced our understanding of the risk factors contributing to RF and might lead us clinical decision to improve the outcome of immediate IBBR.

## Introduction

Breast cancer is the most common carcinoma among female in China and 70% of patients are younger than 60 years old [1]. Along with significant improvements in comprehensive treatment and multidisciplinary team approach, the mortality rate of breast cancer is decreasing [2]. Moreover, as the development of economy and the improvement of living conditions in China, patients’ demand for better body image and quality of life have also been increasing, in especial of younger patients. Breast reconstruction following mastectomy can help patients modify local deformity, improve cosmetic outcome, and thus increase self-confidence and satisfaction in social and psychological aspects [3].

Immediate breast reconstruction has been shown satisfactory aesthetic results and overall psychological well-being compared with delayed breast reconstruction [4]. Moreover, recent studies supported oncologic safety in patients who received immediate breast reconstruction following total mastectomy [5,6]. Similar with the United States, implant-based breast reconstruction (IBBR) represents the most prevalent form of breast reconstruction in China, partly due to low damage without donor site sacrifice and thus postoperative recovery is fast, furthermore, the operation takes a relative shorter learning curve, which is favored by more doctors and patients [7]. However, IBBR is often associated with higher incidence of complications, including infection, implant exposure, capsular contracture, and malposition. Adjuvant therapy, such as chemotherapy and post-mastectomy radiation therapy (PMRT), at further increase the risk of complications [8].

Many tailored strategies, such as two-stage procedure, appropriate incision design, coverage of prosthesis surface, and optimal time of adjuvant therapy were explored for each individual patient to achieve a successful and satisfied reconstruction. However, reconstruction failure (RF), defined as tissue expander (TE) or implant removal due to complications, still has a high rate of 17.6% [9]. Once RF occurs, it is not only a financial loss, but also a frustrate both for surgeons and patients. Surgeons may experience a feeling of guilty and their self-confidence may be undermined, while patients may experience several physical and psychosocial impacts [10]. However, many controversies still exist focusing on risk factors affecting reconstruction, such as one- or two-stage prosthesis breast reconstruction, the impact of adjuvant therapy on reconstructed breast [11]. Thus, the purpose of present study is to evaluate risk factors of RF in immediate IBBR procedure from patient characteristics, surgical techniques, and adjuvant therapies aspects, and establish a nomogram model to predict individual RF risk for immediate IBBR to lead clinical decision making.

## Methods

### Study design and patients enrollment

Breast cancer patients underwent breast reconstruction in our single center from January 2011 to December 2019 were retrospectively reviewed following the approval of Zhejiang Cancer Hospital Clinical Research Ethics Committee (IRB-2020-327). Eligible criteria were as follows: (I) Female patients and histologically confirmed with in situ or invasive breast cancer in primary or post neoadjuvant chemotherapy (NACT); (II) Patients underwent immediate IBBR including one- or two-stage procedure: inserting a permanent implant or TE (classified as stage I operation), exchanging of TE for a permanent implant or followed by flap reconstruction (classified as stage II operation). For data analysis in stage I operation, patients were excluded with any of the following: (I) Delayed breast reconstruction; (II) Immediate autologous breast reconstruction (ABR); (III) Contralateral prophylactic mastectomy (CPM) followed by reconstruction; (IV) Ipsilateral breast radiotherapy history; (V) Patients progressed local recurrence or/and distant metastasis. For data analysis in stage II operation, patient cohort was further excluded with any of the following: (I) Exchange of TE for ABR; (II) TE with no subsequent exchange; (III) TE removed with patients’ requirements (Fig 1).

**Fig 1.**
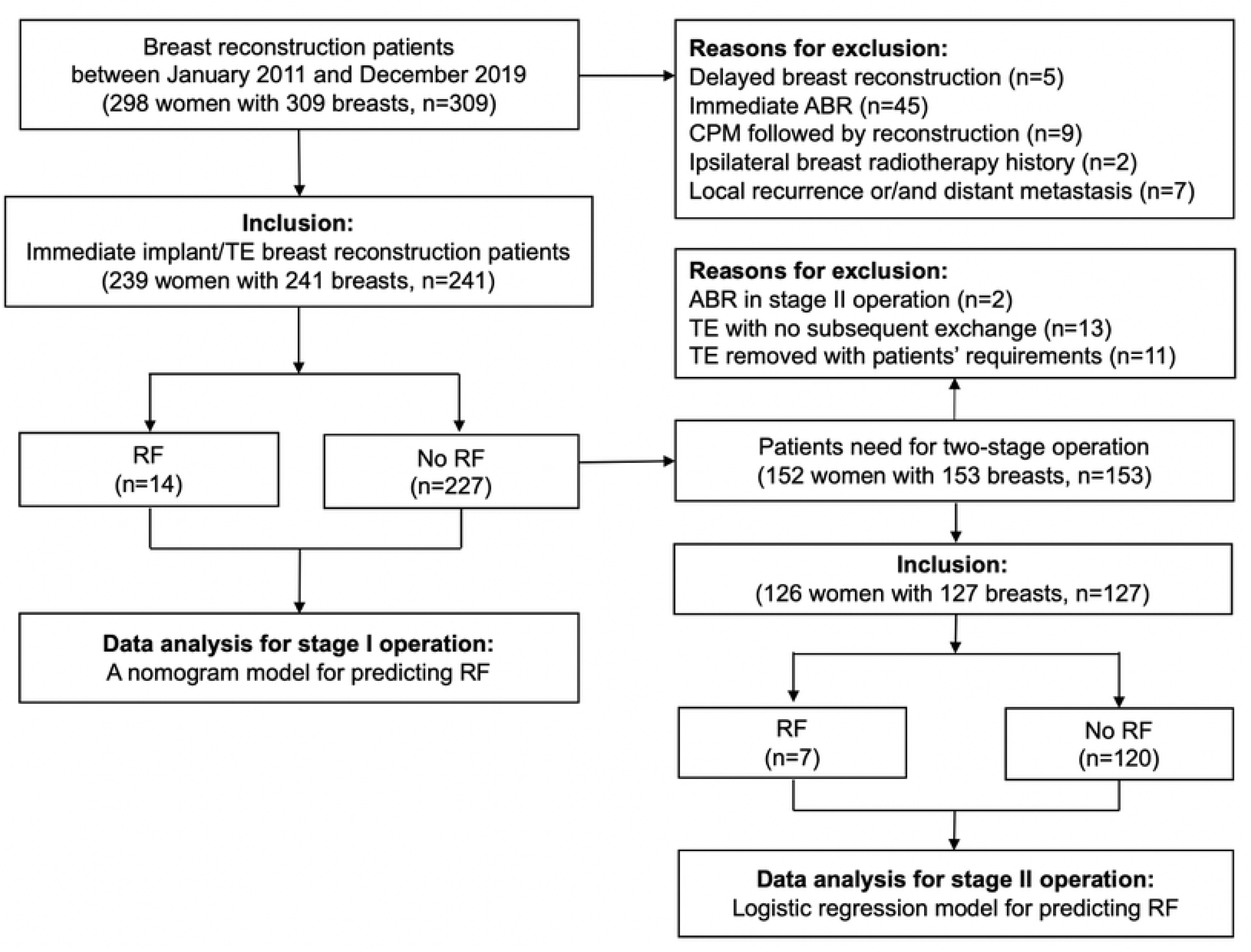
Flow chart for study design and patients enrollment. TE, tissue expander; RF, reconstruction failure; ABR, autologous breast reconstruction; CPM, contralateral prophylactic mastectomy.

### Data collection

Patient characteristics including age, body mass index (BMI; calculated as weight in kilograms divided by height in meters squared), smoking status, comorbidities (diabetes, hypertension), pathological TNM stage according to 8^th^ edition of the AJCC TNM staging system [12] were recorded. Surgical techniques in stage I operation such as skin-sparing mastectomy (SSM) or nipple-sparing mastectomy (NSM), incision types, sentinel lymph node biopsy (SLNB) or axillary lymph node dissection (ALND), volume of implant or TE inflation at surgery, mesh use, device coverage were collected. Adjuvant therapies details such as NACT, adjuvant chemotherapy (ACT), PMRT, perioperative antibiotics were recorded. Incisions for stage II operation were recorded. The duration days from stage I operation to ACT or PMRT, and months from PMRT to stage II operation were collected.

### Surgical techniques

Patients following NACT were respected to receive surgery at least 2 weeks from last chemotherapy. For patients clinically suspected or/and pathologically confirmed of nipple involved, transverse ellipse incision to remove the nipple-areola complex was chosen for performing SSM, while lateral transverse with or without semicircle periareolar, or inframammary fold were common incisions for NSM. In the subpectoral group, implant/TE would be inserted into a pocket combined of pectoralis major and either a mesh or the serratus anterior muscle, whereas in the prepectoral group, partial of latissimus dorsi or skin envelop covered implant/TE. In immediate two-stage IBBR, TE was filled with a variable volume of saline in stage I operation. Multidisciplinary treatment was made for ACT or/and PMRT according to final pathology. For immediate two-stage IBBR, TE was inflated by saline injection 4 weeks later after stage I operation, usually 50-80ml every 2 weeks. TE was required fully expansion, usually extra 20% volume lager than intended implant, before PMRT. Stage II operation was performed at least 3 months from stage I operation, and at least 6 months from radiotherapy. In stage II, some surgeons explored another incision to lower the RF rate. Perioperative antibiotic prophylaxis would be given for 48 hours in immediate IBBR while second-generation cephalosporins were preferred and lincosamides may be another choice for allergic patients.

### Statistical analysis

For analyzing predictors of RF in stage I operation, the volume of implant/TE which inserting to the prepared pocket was calculated as volume percentage according to mastectomy specimen weight and then was divided into two groups according to maximizing the Youden index by receiver operating characteristic (ROC) curve analysis (13). Then enrolled patients were randomly divided into training set and validation set (7:3) by using createDataPartition function in caret package. Continuous variables were analyzed using Student’s t test or Mann-Whitney U test and were reported as mean ± standard deviation (SD). Categorical variables were analyzed using Chi-square test or Fisher’s exact test. Logistic regression analysis was performed and potential prognostic factors (*p*<0.1) by univariate analysis further entered multivariate analysis to identify independent predictors of RF. Then a nomogram for predicting RF in stage I was established based on the logistic model using rms package. The nomogram was quantified by discrimination and calibration both in training set and validation set. For analyzing predictors of RF in stage II operation, logistic regression (Method=Enter) was used for multivariate analysis to identify independent predictors. Statistical analysis was performed using SPSS software (version 25.0; IBM Corporation, Armonk, NY, USA) and R software (version 4.0.3; https://www.r-project.org/). All reported *p* values were two-sided, and *p*<0.05 was considered statistically significant.

## Results

A total of 298 consecutive breast cancer patients (309 breasts) receiving breast reconstruction surgery from January 2011 to December 2019 in our single center were retrospectively reviewed and finally 239 patients (241 breasts) who met the inclusion criteria were enrolled for the study cohort. The median follow-up time since breast reconstruction surgery was 48 months (21-123) months. During nine years in our single center, both the use of two-stage operation and NSM increased (Fig 2A-2B), and rates of RF per year remain stable lower than 10% (Fig 2C). Overall, 14 (5.81%) and 7 (5.51%) reconstructive breasts experienced failure in stage I and II operation, respectively.

**Fig 2.**
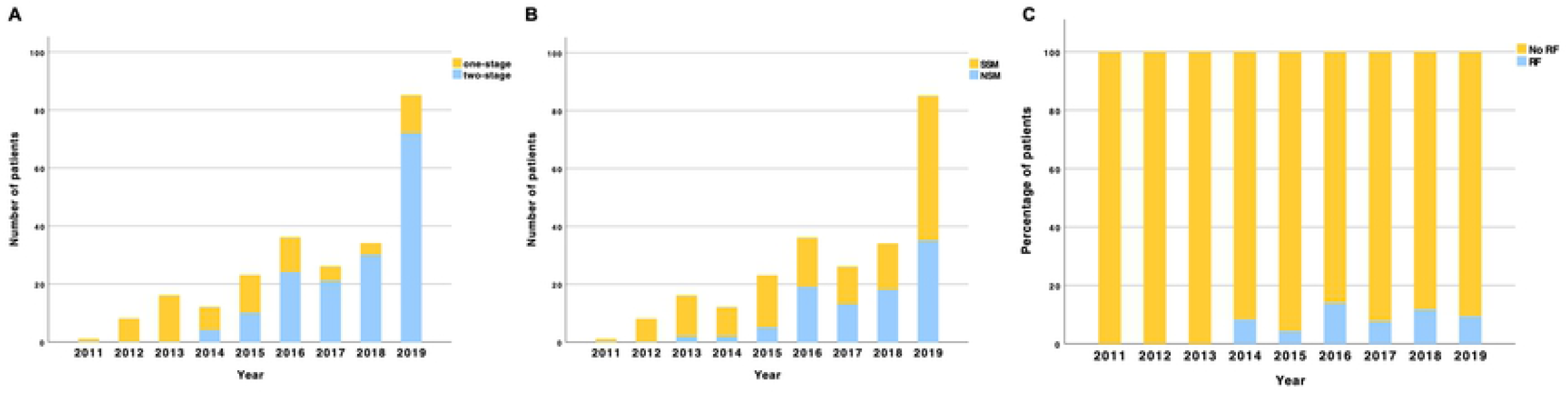
(A) Trends of one- or two-stage operation in immediate IBBR. (B) Trends of SSM and NSM in immediate IBBR. (C) RF rate of immediate IBBR per year in our single center. IBBR, implant-based breast reconstruction; NSM, nipple-sparing mastectomy; SSM, skin-sparing mastectomy; RF, reconstruction failure.

### Characteristics of study cohort and treatment

To establish a nomogram for predicting RF, the study cohort was randomly (7:3) divided into training cohort (169 breasts) and validation cohort (72 breasts). The characteristics of patients, surgical techniques, and adjuvant therapies of the training and validation cohorts were summarized in Table 1. All patients had no history of smoking (present or former) and no patient had comorbidity of diabetes or hypertension. The optimal cut-off value for implant/TE volume by ROC curve analysis was 86.30%. 34 (14.11%) implant/TEs suffered immediate PMRT (all of them underwent surgery after NACT and the time between surgery to PMRT ranged from 1-3 months) and one implant lost. 33 implant/TEs experienced radiotherapy after ACT, usually 4-7 months later from surgery and no RF occurred.

**Table 1.** Characteristics for the training and validation cohorts of 241 breasts underwent stage I operation of immediate implant-based breast reconstruction

| Characteristics | Overall cohort<br>(n=241), n (%) | Training cohort<br>(n =169), n (%) | Validation cohort<br>(n = 72), n (%) | p value |
| --- | --- | --- | --- | --- |
| Age (mean ± SD), years | 39.81 ± 7.77 | 39.30 ± 7.69 | 41.03 ± 7.86 | 0.932 |
| BMI (mean ± SD), kg/m<sup>2</sup> | 21.99 ± 2.48 | 21.94 ± 2.35 | 22.11 ± 2.75 | 0.212 |
| yp/pTNM stage |  |  |  |  |
| 0 | 30 (12.45) | 22 (13.02) | 8 (11.11) | 0.967 |
| I | 102 (42.32) | 72 (42.60) | 30 (41.67) |  |
| II | 81 (33.61) | 56 (33.14) | 25 (34.72) |  |
| III | 28 (11.62) | 19 (11.24) | 9 (12.50) |  |
| Mastectomy type |  |  |  |  |
| SSM | 147 (61.00) | 102 (60.36) | 45 (62.50) | 0.775 |
| NSM | 94 (39.00) | 67 (39.64) | 27 (37.50) |  |
| Incision |  |  |  |  |
| Without periareolar | 233 (96.68) | 162 (69.53) | 71 (30.47) | 0.442 |
| Periareolar | 8 (3.32) | 7 (87.50) | 1 (12.50) |  |
| Lymph node management |  |  |  |  |
| SLNB | 149 (61.83) | 106 (71.14) | 43 (28.86) |  |
| ALND | 92 (38.17) | 63 (68.48) | 29 (31.52) | 0.667 |
| Implant/TE volume |  |  |  |  |
| ≤86.30% | 168 (69.71) | 121 (71.60) | 47 (65.28) | 0.360 |
| > 86.30% | 73 (30.29) | 48 (28.40) | 25 (34.72) |  |
| Mesh use |  |  |  |  |
| No | 178 (73.86) | 124 (73.37) | 54 (75.00) | 0.873 |
| Yes | 63 (26.14) | 45 (26.63) | 18 (25.00) |  |
| Covering of pocket |  |  |  |  |
| Latissimus dorsi | 48 (19.92) | 28 (16.57) | 20 (27.78) | 0.131 |
| Pectoralis major muscle | 182 (75.51) | 133 (78.70) | 49 (68.06) |  |
| Skin | 11 (4.57) | 8 (4.73) | 3 (4.17) |  |
| NACT |  |  |  |  |
| No | 204 (84.65) | 142 (84.02) | 62 (86.11) | 0.845 |
| Yes | 37 (15.35) | 27 (15.98) | 10 (13.89) |  |
| ACT |  |  |  |  |
| No | 103 (42.74) | 71 (42.01) | 32 (44.44) | 0.777 |
| Yes | 138 (57.26) | 98 (57.99) | 40 (55.56) |  |
| Immediate PMRT |  |  |  |  |
| No | 207 (85.89) | 143 (84.62) | 64 (88.89) | 0.426 |
| Yes | 34 (14.11) | 26 (15.38) | 8 (11.11) |  |
| Perioperative antibiotics |  |  |  |  |
| Lincosamides | 26 (10.79) | 17 (10.06) | 9 (12.50) | 0.651 |
| Cephalosporins | 215 (89.21) | 152 (89.94) | 63 (87.50) |  |
| RF |  |  |  |  |
| No | 227 (94.19) | 159 (94.08) | 68 (94.44) | 1.000 |
| Yes | 14 (5.81) | 10 (5.92) | 4 (5.56) |  |

SD, standard deviation; BMI, body mass index; SSM, skin-sparing mastectomy; NSM, nipple-sparing mastectomy; SLNB, sentinel lymph node biopsy; ALND, axillary lymph node dissection; TE, tissue expander; NACT, neoadjuvant chemotherapy; ACT adjuvant chemotherapy; PMRT, post-mastectomy radiation therapy; RF, reconstruction failure.

### Predictors of reconstruction failure in stage I operation

Analysis of 169 breasts in the training cohort revealed that 10 breasts experienced RF, then characteristics between RF and no RF groups were compared (Table 2). Logistic regression analysis was performed and variables in case of zero count were excluded (variables of incision, skin covering pocket and NACT) in the training cohort. Univariate analysis showed that BMI, lymph node management, implant/TE volume, mesh use and ACT were possible factors associated with RF (p<0.1). Further multivariate analysis revealed that BMI [odds ratio (OR)=1.569, 95% confidence interval (CI): 1.087-2.263, P=0.016], implant/TE volume>86.30% (OR=5.711, 95% CI: 1.067-30.583, P=0.042), ACT (OR=30.094, 95% CI: 2.030-446.084, P=0.013) were independent risk factors for RF (Table 3).

**Table 2.**
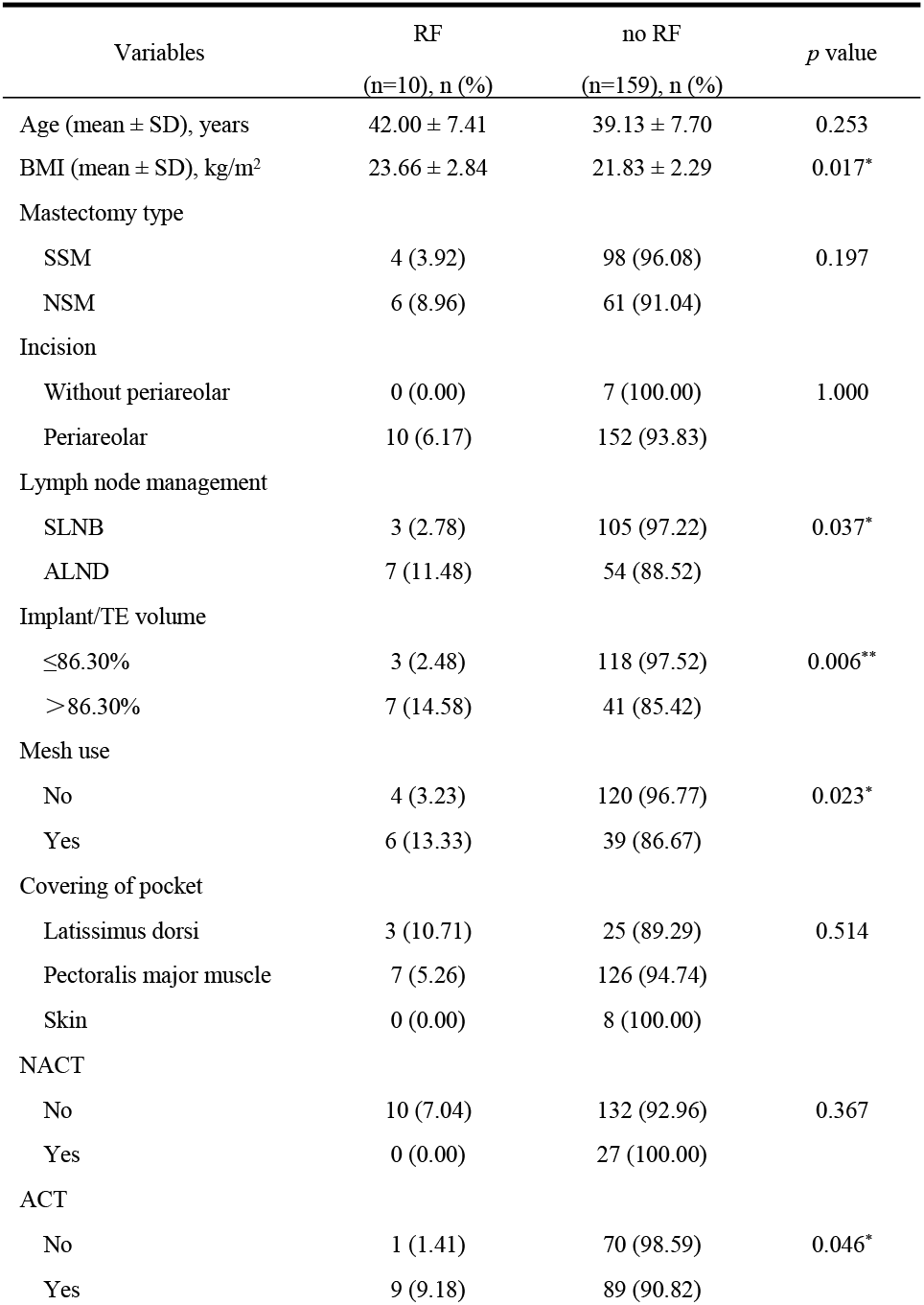
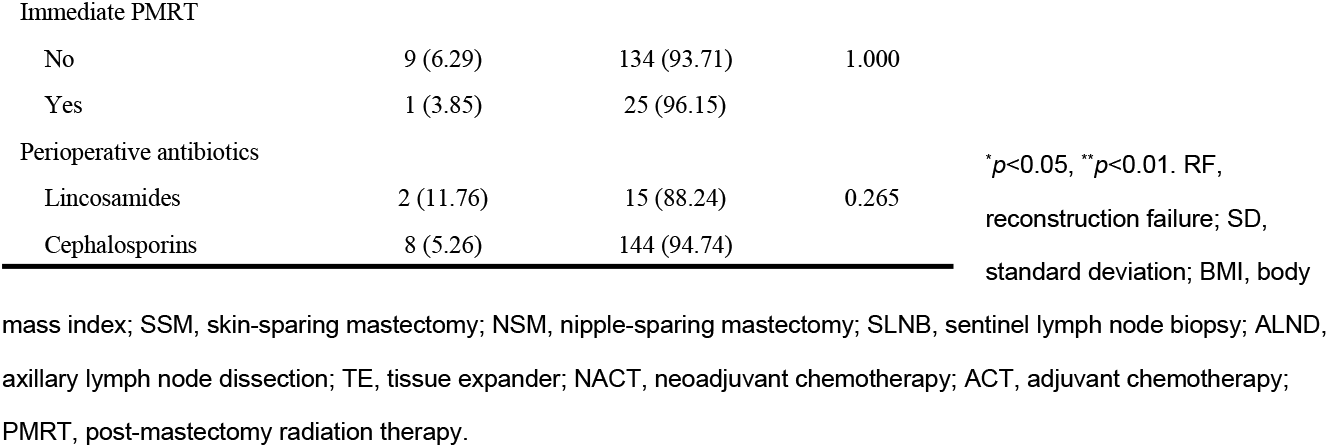
Comparisons of characteristics in RF and no RF groups of the training cohort in stage I operation

| Variables | RF<br>(n=10), n (%) | no RF<br>(n=159), n (%) | *p* value |
| --- | --- | --- | --- |
| Age (mean ± SD), years | 42.00 ± 7.41 | 39.13 ± 7.70 | 0.253 |
| BMI (mean ± SD), kg/m<sup>2</sup> | 23.66 ± 2.84 | 21.83 ± 2.29 | 0.017* |
| Mastectomy type |  |  |  |
| SSM | 4 (3.92) | 98 (96.08) | 0.197 |
| NSM | 6 (8.96) | 61 (91.04) |  |
| Incision |  |  |  |
| Without periareolar | 0 (0.00) | 7 (100.00) | 1.000 |
| Periareolar | 10 (6.17) | 152 (93.83) |  |
| Lymph node management |  |  |  |
| SLNB | 3 (2.78) | 105 (97.22) | 0.037* |
| ALND | 7 (11.48) | 54 (88.52) |  |
| Implant/TE volume |  |  |  |
| $\leq 86.30\%$ | 3 (2.48) | 118 (97.52) | 0.006** |
| $> 86.30\%$ | 7 (14.58) | 41 (85.42) | |
| Mesh use |  |  |  |
| No | 4 (3.23) | 120 (96.77) | 0.023* |
| Yes | 6 (13.33) | 39 (86.67) |  |
| Covering of pocket |  |  |  |
| Latissimus dorsi | 3 (10.71) | 25 (89.29) | 0.514 |
| Pectoralis major muscle | 7 (5.26) | 126 (94.74) |  |
| Skin | 0 (0.00) | 8 (100.00) |  |
| NACT |  |  |  |
| No | 10 (7.04) | 132 (92.96) | 0.367 |
| Yes | 0 (0.00) | 27 (100.00) |  |
| ACT |  |  |  |
| No | 1 (1.41) | 70 (98.59) | 0.046* |
| Yes | 9 (9.18) | 89 (90.82) |  |
| Immediate PMRT |  |  |  |
| No | 9 (6.29) | 134 (93.71) | 1.000 |
| Yes | 1 (3.85) | 25 (96.15) |  |
| Perioperative antibiotics |  |  |  |
| Lincosamides | 2 (11.76) | 15 (88.24) | 0.265 |
| Cephalosporins | 8 (5.26) | 144 (94.74) |  |

\* $p < 0.05$ , \*\* $p < 0.01$ . RF, reconstruction failure; SD, standard deviation; BMI, body
mass index; SSM, skin-sparing mastectomy; NSM, nipple-sparing mastectomy; SLNB, sentinel lymph node biopsy; ALND, axillary lymph node dissection; TE, tissue expander; NACT, neoadjuvant chemotherapy; ACT, adjuvant chemotherapy; PMRT, post-mastectomy radiation therapy.

**Table 3.** Logistic regression analysis of reconstruction failure in the training cohort (169 breasts)

| Variables | Univariate Analysis |  | Multivariate Analysis |  |
| --- | --- | --- | --- | --- |
|  | OR (95% CI) | *p* value | OR (95% CI) | *p* value |
| Age, years | 1.052 (0.964-1.149) | 0.254 |  |  |
| BMI, kg/m<sup>2</sup> | 1.388 (1.051-1.834) | 0.021* | 1.569 (1.087-2.263) | 0.016* |
| Mastectomy type |  |  |  |  |
| SSM | 1.000 |  |  |  |
| NSM | 2.410 (0.654-8.886) | 0.186 |  |  |
| Lymph node management |  |  |  |  |
| SLNB | 1.000 |  |  |  |
| ALND | 4.537 (1.128-18.248) | 0.033* | 4.876 (0.966-24.617) | 0.055 |
| Implant/TE volume |  |  |  |  |
| ≤86.30 | 1.000 |  |  |  |
| >86.30 | 6.715 (1.659-27.189) | 0.008** | 5.711 (1.067-30.583) | 0.042* |
| Mesh use |  |  |  |  |
| No | 1.000 |  |  |  |
| Yes | 4.615 (1.238-17.204) | 0.023* | 3.510 (0.682-18.073) | 0.133 |
| Position |  |  |  |  |
| Latissimus dorsi | 1.000 |  |  |  |
| Pectoralis major muscle | 0.463 (0.112-1.913) | 0.287 |  |  |
| ACT |  |  |  |  |
| No | 1.000 |  |  |  |
| Yes | 7.079 (0.876-57.206) | 0.066 | 30.094 (2.030-446.084) | 0.013* |
| Immediate PMRT |  |  |  |  |
| No | 1.000 |  |  |  |
| Yes | 0.596 (0.072-4.911) | 0.630 |  |  |
| Perioperative antibiotics |  |  |  |  |
| Lincosamides | 1.000 |  |  |  |
| Cephalosporins | 0.417 (0.081-2.144) | 0.295 |  |  |

\* $p < 0.05$ , \*\* $p < 0.01$ . OR, odds ratio; BMI, body mass index; SSM, skin-sparing mastectomy; NSM, nipple-sparing mastectomy; SLNB, sentinel lymph node biopsy; ALND, axillary lymph node dissection; TE, tissue expander; ACT, adjuvant chemotherapy; PMRT, post-mastectomy radiation therapy.

### Nomogram establishment and validation

A nomogram was established based on five variables, including BMI, lymph node management, implant/TE volume, mesh use and ACT to predict the RF rate in stage I operation. The points for the five factors were summed up to calculate the probability of RF (Fig 3). ROC curves and calibration curves were created based on the training and validation cohorts for internal and external validation, respectively (Fig 4). The ROC curve of the nomogram revealed area under the curve (AUC) was 0.920 (95% CI: 0.844-0.995) (Fig 4A) and external validation revealed AUC of 0.930 (Fig 4B), showing a satisfactory discriminative ability of the nomogram. The calibration curve based on training cohort with a bootstrap resampling frequency of 1000 and a Hosmer-Lemeshow Chi square value of 8.946 (*p*=0.347), showed a goodness of fit. Additionally, brier score was 0.040, showing a good calibration ability of the nomogram. Calibration curves of training (Fig 4C) and validation (Fig 4D) cohorts showed an acceptable consistency between the actual observation and logistic model prediction.

**Fig 3.**
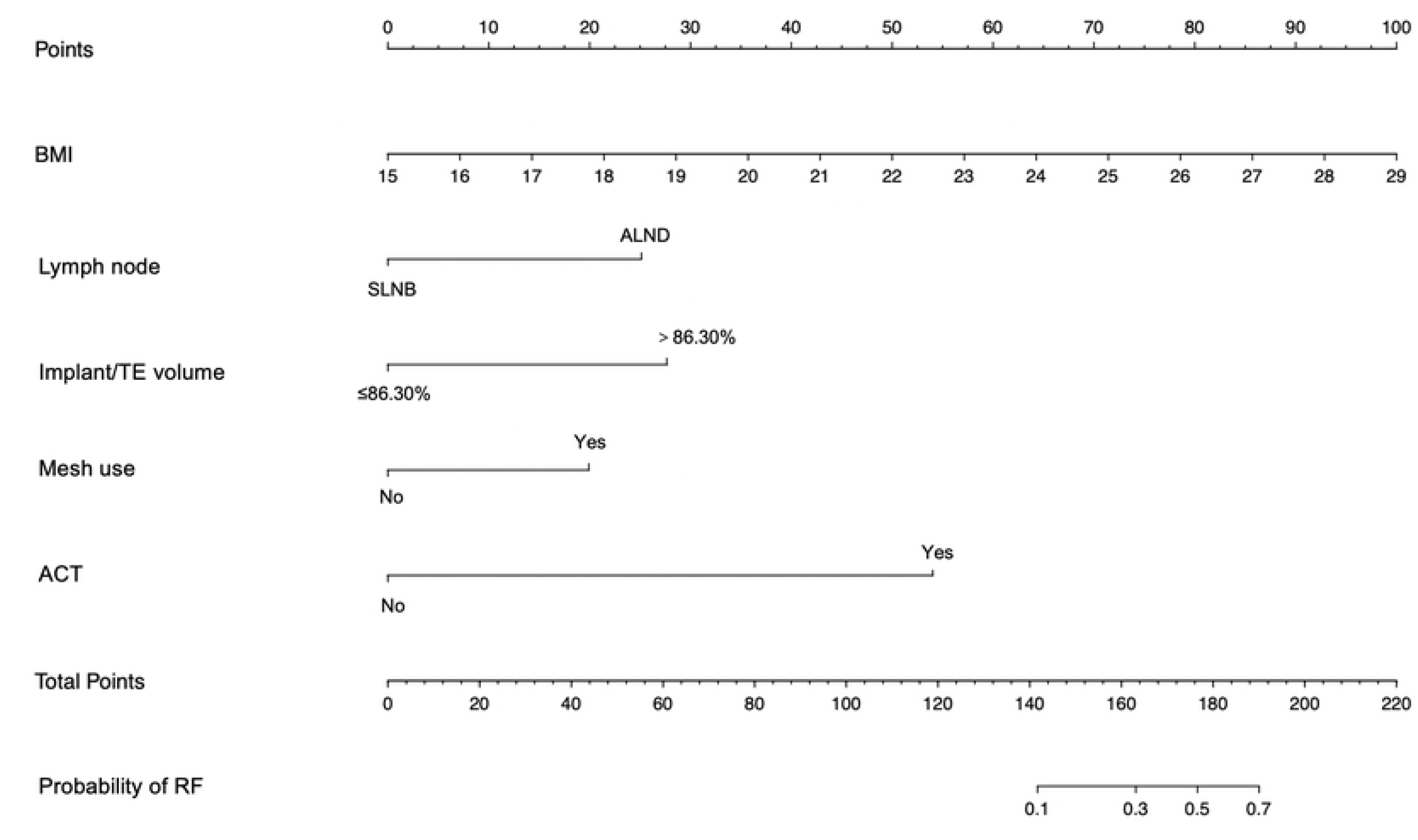
A nomogram to predict the RF rate in stage I operation. BMI, body mass index; TE, tissue expander; SLNB, sentinel lymph node biopsy; ALND, axillary lymph node dissection; ACT, adjuvant chemotherapy; RF, reconstruction failure.

**Fig 4.**
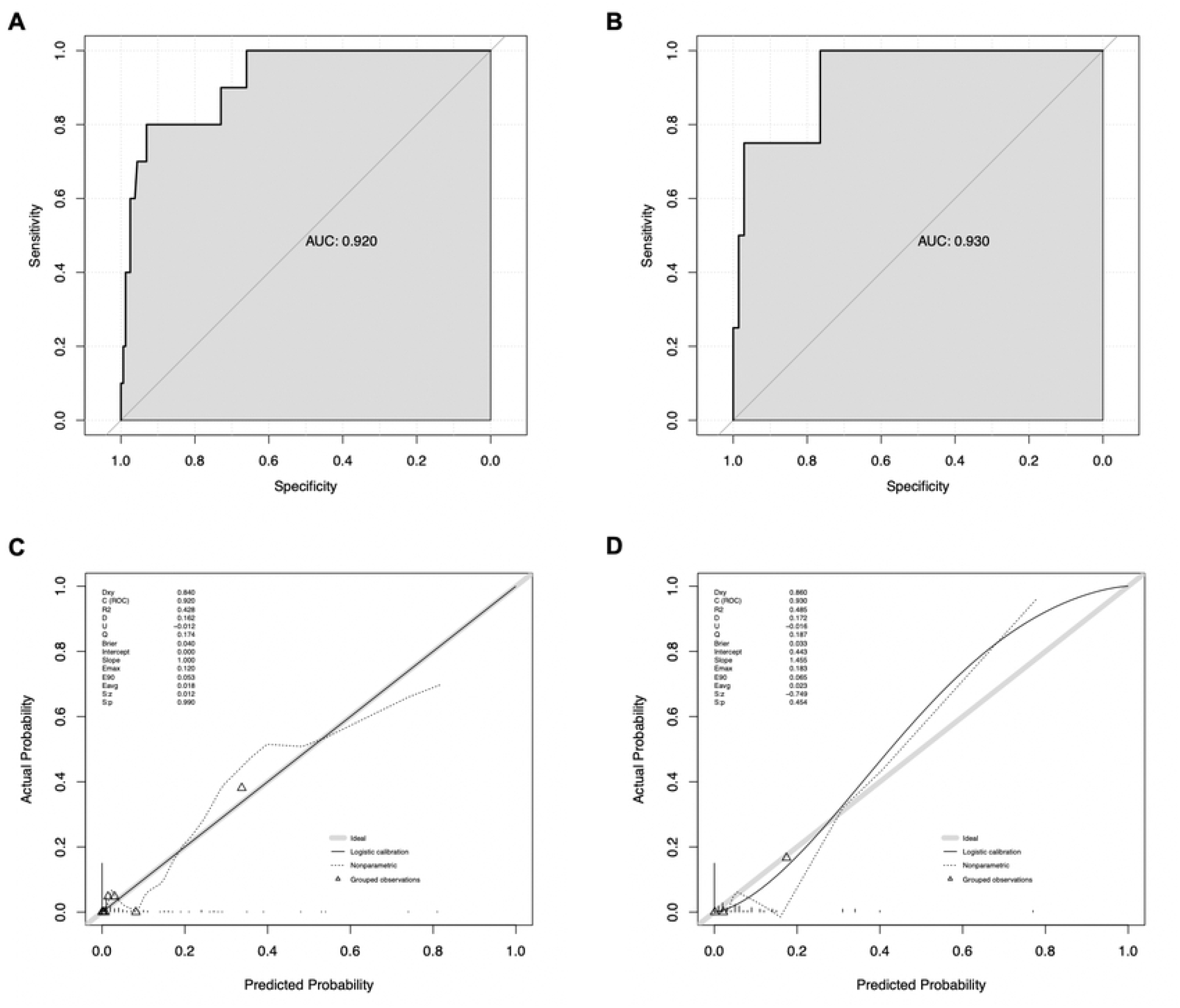
ROC curves with an AUC of 0.920 derived from the training cohort (A) and 0.930 derived from the validation cohort (B) demonstrated the discriminatory ability of the nomogram. The calibration curves for the nomogram as the internal validation group derived from the training cohort (C) and external calibration group derived from the validation cohort (D). AUC; area under the curves; ROC, receiver operating characteristic.

### Predictors of reconstruction failure in stage II operation

A total of 127 breasts met the inclusion criteria for analysis in stage II operation. There were 7 (5.51%) reconstructive breasts experienced failure in stage II operation. While comparing characteristics with and without RF, there was no significant difference in age, BMI, incision, pocket coverage, radiotherapy history or perioperative antibiotics. RF rate was significantly higher in the mesh use group (17.65% vs 1.07%; *p*=0.001). Multivariate analysis demonstrated that mesh use was the independent predictor of RF stage II operation (OR=47.326, 95% CI: 3.11-720.081, *p*=0.005) (Table 4).

**Table 4.**
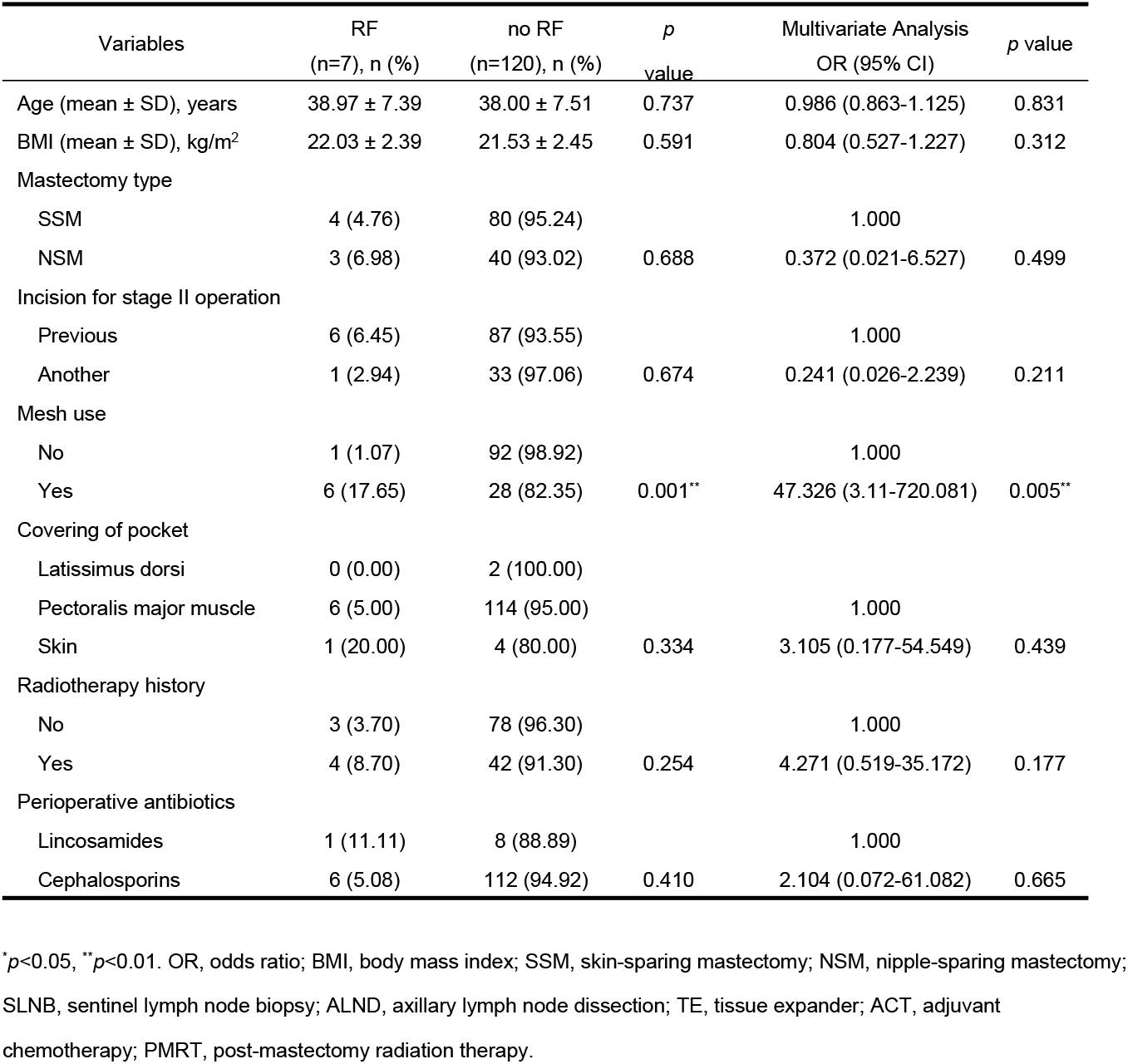
Comparison of variables between RF with no RF groups and multivariate logistic regression model for predicting RF in stage II operation

## Discussion

With the rising trend of immediate IBBR surgery over past nine years in our single center, the overall reconstruction failure rates were 5.81% and 5.51% in stage I and II operation, respectively, which were still acceptable and relatively lower compared with previous literatures (5.6%-22.9%) [14,15]. Each stage of immediate IBBR was separated with its own risks, so risks of RF were compared for each stage separately in present study. A nomogram was established to predict RF rate in stage I operation and a logistic regression model was established to predict RF rate in stage II operation, separately. The nomogram was established based on five variables, including BMI, lymph node management, implant/TE volume, mesh use and ACT. Among these variables, higher BMI, implant/TE volume>86.30% and ACT were independent risk factors for RF in stage I operation. During stage II operation, a logistic regression model was established, and mesh use was demonstrated as the independent predictor of RF. To our knowledge, this is the first clinical study to use nomogram to develop a prediction model of RF rate for immediate IBBR. Additionally, with an AUC of 0.920 (95% CI: 0.844-0.995) and an AUC of 0.930 via external validation, the model exhibited satisfied predict performance.

Old age and higher BMI were well-established risk factors for RF in immediate IBBR from patient aspect [16]. Furthermore, many studies indicated obesity was associated with RF in both implant-based and autologous breast reconstruction [17,18]. The mean age and BMI in present study were 39.81 years and 21.99 kg/m^2^, respectively, indicating patients underwent immediate IBBR in this study were mostly young and normal in weight. Age was not a risk factor in our study both in stage I and II operations. In contrast, higher BMI was be found significantly associated with RF in stage I operation, with an OR of 1.569 compared with relative lower BMI individuals. However, BMI was not a risk factor in stage II operation. Some studies have proposed a theory to explain this result, because higher BMI women might have larger breasts, which led to longer operative time, longer mastectomy flaps and more ischemia flaps. While these high-risk patients might not progress to stage II operation due to removal of TE in stage I operation [8,19].

Skin-sparing mastectomy was a widespread technique in breast reconstruction surgery, which preserves native skin envelope and inframammary fold [20]. By contrast, nipple-sparing mastectomy was an improved satisfaction surgical technique with proven oncological safety in appropriately selected patients [21,22]. However, different incision locations for NSM procedure were associated with different rates of NAC necrosis. Previous literatures demonstrated that periareolar incisions had a higher NAC necrosis rate which might result in RF [23]. 39.00% of breasts in present study retained NAC, and 8 [3.32%] breasts received periareolar incisions. Nevertheless, mastectomy type and incision were not risk factors in our study in stage I operation.

There were significant differences in axillary lymph node management between RF and no RF groups during stage I operation. ALND was associated with higher risk of RF in univariate analysis, however, it was not an independent risk factor by multivariate analysis. Removal of axillary lymph nodes affects lymph drainage and wound healing, which results in postoperative complication such as flap lymphedema, wound infection, and delayed healing [24]. Some studies demonstrated that seroma formation was the most likely reason for wound infection after ALND and minimizing dead space through fixation axillary flaps with underlying muscles can lower the incidence of seroma [25,26]. We might try this method for improvement in future surgery procedure.

Perfusion of the flap was closely associated with complication of delayed wound healing in IBBR. Yang et al. quantified tension-perfusion relationship and proved that the flap perfusion deteriorated as skin tension increased from larger implant [27]. Permanent implant in one-stage breast reconstruction, whose volume cannot be manipulated, causes greater tension on the skin envelope. In case of SSM techniques, whose skin envelope was reduced due to NAC dissection, will inherently create further stress to skin envelop [11]. On the contrary, the volume of TE can be manipulated by inflation or deflation. Our study indicated that 86.30% of the mastectomy specimen weight was the cutoff of implant/TE volume when inserting to prepared pocket in one-stage immediate IBBR or in stage I operation of two-stage immediate IBBR. Moreover, implant/TE volume>86.30% was independent risk factor for RF in stage I operation. Recently, some studies recommended indocyanine green fluorescence to assess the perfusion of flap during surgery, and volume of TE can be modulated to ensure the skin perfusion and the skin with ischemic condition can be removed to reduce complications [28]. We tried indocyanine green fluorescence assessment in several cases and indeed found relatively accurate evaluation of flap perfusion, but unfortunately, sample size was too small for us to gather and analyze data.

The coverage of implant/TE would be skin, total muscular, or muscular combined with a mesh in our study. The covering of pocket was not associated with RF rate both in stage I and II operation. Total muscular coverage provides a protective pocket for implant/TE when flap necrosis or wound disruption [29]. Mesh was generally placed and sutured with pectoralis major muscle edges to support and cover prosthesis. The mesh has many advantages such as reducing muscle sacrifice, reducing prosthesis migration, improving better pole, and resulting in natural inframammary fold, specifically in cases of larger size of breast or insufficient muscle coverage [11]. Nevertheless, mesh use significantly increased the risk of complications compared with total muscular coverage in IBBR [30]. Titanium-coated polypropylene mesh, which was approved for use in 2008 and the implant loss rate were 8.7% in previous report [31], was popularly used in our center. Our study showed mesh use was associated with higher risk of RF in univariate analysis although it was not an independent risk factor by multivariate analysis in stage I operation. Furthermore, mesh use was the independent risk factor for RF in stage II operation.

Chemotherapeutic agents and irradiation have been proven to reduce local recurrence and improve patient survival outcome in advanced breast cancer [32]. However, many studies demonstrated that adjuvant chemotherapy was associated with cytotoxicity effect and immunosuppression which caused wound infection, delayed healing [33]. Similarly, PMRT on prosthesis might increase complication and compromise aesthetic outcome [34]. One systematic review showed that RF rate was significantly lower when radiotherapy was given after stage II (5.6%, radiotherapy to implant) compared to radiotherapy given after stage I (22.9%, radiotherapy to TE). However, it was unclear how many implants were lost during chemotherapy period of treatment following stage I without experiencing radiotherapy [14]. In stage I operation of our study, RF rate was 0.97% (1/103), 8.70% (12/138), 2.94% (1/34) for reconstructive breasts with no ACT, with ACT, with immediate PMRT, respectively. Meanwhile, radiotherapy history, which contained immediate PMRT and radiotherapy after ACT, was not associated with RF in stage II operation. Obviously, radiotherapy was not the only culprit in IBBR. Nevertheless, ACT was considered as an independent risk factor for RF in stage I operation.

Two-stage technique of breast reconstruction is the most commonly form of immediate IBBR today [35]. Advocates for two-stage operation proposed the ability of revision on radiation effect and symmetries in stage II operation to obtain more satisfactory appearance, and because complications rate was relatively lower when compared with single-stage reconstruction [11]. Some surgeons in our center explored to choose another incision to lower RF rate in stage II operation, especially for patients who had radiotherapy history, for the theory that poor flap perfusion and delayed healing after radiotherapy. However, the incision was not the predict factor of RF in stage II operation. Many literatures demonstrated that 24 hours of antibiotic prophylaxis was warranted [36] and infectious disease guidelines recommended cephalosporins against Gram-positive organisms are appropriate for nonallergic [37]. All patients in our study received antibiotic prophylaxis for 48 hours during the perioperative period, second-generation cephalosporins for nonallergic patients and lincosamides for others. Moreover, the perioperative antibiotic was not the predictor for RF.

Although the nomogram model created in our study demonstrated satisfactory performance, we still acknowledged some limitations. Firstly, as the data was retrospectively reviewed, some important details were unavailable such as patients’ satisfaction score of reconstructed breasts, Baker grading following radiotherapy. Secondly, multiple breast and plastic surgeons might have variative learning curve of reconstructive surgery technique, which also affected the outcome and complication rate. Thirdly, the number of patients is relatively smaller due to the low incidence rate of events. Additionally, only logistic regression model is suitable due to the limited sample and variables in analysis of stage II operation, which resulted in insufficient data for validation. Therefore, a multicenter perspective study with more data available is necessary to overcome these limitations.

## CONCLUSION

This is the first application of nomogram to develop a prediction model of RF rate for immediate IBBR, which demonstrated higher BMI, implant/TE volume>86.30% and ACT were independent risk factors for RF in stage I operation and the model exhibited satisfied predictive ability. Then a logistic regression model was established, and mesh use was independent predictor of RF in stage II operation. The nomogram created in this study is an easy-to-use tool for clinicians to predict RF rate in immediate one-stage IBBR or stage I operation of immediate two-stage IBBR. Furthermore, these findings enhanced our understanding of the risk factors contributing to RF and might lead us clinical decision to improve the outcome of immediate IBBR.

## Data Availability

All relevant data are within the manuscript and its Supporting Information files.

## Acknowledgments

This work is funded by grants from the Medical Health Science and Technology Project of Zhejiang Provincial Health Commission (2021RC045).

